# Spatial Patterns and Determinants of Climate Change Awareness and Implications for Humanitarian Health Response in Nigeria: A Cross-Sectional Analysis of a Nationally Representative Survey

**DOI:** 10.64898/2026.05.12.26352814

**Authors:** Abolaji Moses Ogunetimoju, Oluwafemi Lawal Bisiriyu, Kehinde Peter Ajewole, Temisola Emmanuel Oyelakin

## Abstract

**Objectives:** To explore the prevalence, spatial aggregation, and demographic correlates of climate change awareness among adults in Nigeria, as well as impacts on humanitarian health preparedness.

**Design:** Nationally representative cross-sectional survey with multivariate logistic regression and Global Moran’s I and LISA techniques of spatial autocorrelation analyses was applied.

**Settings:** All 36 states and the Federal Capital Territory, Nigeria.

**Participants:** 1,600 adults drawn from the Afrobarometer Round 9 nationally representative survey.

**Interventions:** None

**Main Outcome:** Measures Prevalence, spatial aggregation, and demographic correlates of climate change awareness among adults in Nigeria, and impacts on humanitarian health preparedness.

**Results:** Less than one in three Nigerians (30.1%) was aware of climate change, significantly lower than the 65% found in the continent, and education is the most predictive factor, with tertiary-educated Nigerians more than ten times more likely to be aware of climate change than those with no formal education. Most critically, the poor performance in government climate policies is not found in low-awareness states, but in two geographically distinct risk corridors based on a different mechanism and requiring a different policy response.

**Conclusions:** The finding shows that the gap in climate awareness is not a communication problem, it is a structural problem, one that requires a national intervention to reduce and close, but that might not be enough because of educational inequality, gender disparity and geographic marginalization. To prepare the country for humanitarian needs, targeted state-level, gender-responsive programming based on Nigeria’s Climate Change Act 2021 is required, and effective intervention to make adaptation to the health impacts of climate change happen will need to start with triggering awareness into adaptive health action before climate hazards surpass the country’s humanitarian response capacity.

**Registration:** **Not applicable**.

## Introduction

Climate change is one of the twenty first century’s most critical humanitarian issues and Africa is the most vulnerable continent to climate change. This has not spared West Africa and Nigeria, where the threats of higher temperatures, irregular rainfall and desertification in the north, and increased coastal flooding in the south continue to occur and intensify (Intergovernmental Panel on Climate Change [IPCC] 2023). Nigeria, with over 220 million people is one of the most vulnerable countries in Africa, ranking 154th of 185 countries on the Notre Dame Global Adaptation Initiative (ND-GAIN) Climate Vulnerability and Adaptive Capacity (2022) and 2nd place in the global ranking of the Children’s Climate Risk Index (ND-GAIN, 2022; UNICEF, 2021).

The humanitarian health impacts of these hazards are already being observed and are increasing, as the 2022 floods were the worst in ten years, killing 603 people, injuring 2,504, displacing some 1.4 million people in 34 states, and estimated to have cost the economy US$4.2 billion (NEMA, 2023). The subsequent flooding that occurred in 2024 proved more devastating as 1237 people lost their lives and more than 5.2 million Nigerians were affected (NEMA, 2025). To the north, the region has faced the problem of food insecurity due to extended periods of drought, climate-induced displacement, and pastoralist-farmer conflicts, as well as the reduction of Lake Chad (United Nations Security Council, 2024). In addition to these direct physical hazards, climate change is also directly linked to an increase in infectious disease burden in Nigeria, with increases in temperature and irregular rainfall patterns driving increases in vector breeding, while cholera and meningitis outbreaks have also been observed to follow climate-driven seasonal and hydrological trends, further contributing to the country’s disproportionate share of approximately 27% of all malaria cases and nearly 39% of all under-five malaria deaths globally (WHO, 2023; WHO AFRO, 2024).

In the same vein, the level of awareness about climate change among Nigerians continues to be alarmingly low in the face of increasing climate exposure. In Nigeria, Afrobarometer Round 9 survey results from March 2022 found that just 30% of adults had ever heard about climate change, which was one of the lowest rates of the 38 countries surveyed across Africa, and significantly lower than the continent’s average of 51% (Duntoye et al., 2023; Afrobarometer, 2025). This lack of awareness has direct humanitarian implications: there is strong evidence that public awareness is a necessary precondition for both changing individual climate-adaptive behaviors and enhancing community resilience and support for climate policy (van der Linden, 2015; Torsu & Kronke, 2023). Communities cannot make informed adaptive decisions, individuals cannot take protective health actions, and citizens cannot engage in sustainable influence on their climate policy if they are not aware of the driver of the hazards they are facing on a daily basis.

Literature has identified some factors that influence climate change awareness in Africa, for example, Lee et al. (2015) found that educational attainment is the strongest prediction of climate change awareness across 119 countries in the world from their study on Gallup data. Similarly, Simpson et al. (2021) used data from the Afrobarometer survey of Round 7 (2018-2020) to find that education, urbanization, media access and sex were strong, consistent predictors of climate literacy, with Nigeria one of the countries with the lowest rates. Azeez et al. (2024) more recently found that geographic location, information source, and sociodemographic factors were significant factors in determining climate consciousness in 34 countries across Africa. In addition, Oyelakin & Bisiriyu (2026) investigated public perception of climate change based on planetary health. Information on studies that have investigated the spatial clustering of awareness deficits across the states of Nigeria is however, limited and a lack of application of spatial analytical methods has hindered the identification of zones of concentrated climate literacy deficit, which makes it difficult for policy makers and humanitarian practitioners to make proportionate interventions.

This research fills three additional research gaps. Second, national or zonal prevalence estimates, reported elsewhere (Okereke et al., 2025; Agada et al., 2023; Azeez et al., 2024; Lee et al., 2015) are made without considering spatial autocorrelation, that is, whether low-awareness states are grouped together with other low-awareness states. Second, the governance dimension of climate perception, as this relates to the performance of government on climate action and the people they blamed for the lack of mitigation has not gotten enough attention in the peer-reviewed literature. Thirdly, few studies have been conducted on independent predictors of climate change awareness with spatially disaggregated data, particularly from a humanitarian health perspective. When combined, these gaps create a humanitarian health and climate policy community for Nigeria that lacks state-level spatial evidence for guiding targeted humanitarian responses.

This study tackles these gaps by using the Afrobarometer Round 9 Nigeria data and incorporating spatial analytical methods to analyze climate change awareness and perceptions. There are three reasons behind this approach. Nationally representative data allows for statistically sound and spatially relevant findings, first; and state-level disaggregation allows for an informed and effective spatial response, second. Second, the combination of Global Moran’s I and Local Indicators of Spatial Association (LISA) enabled the first spatial analysis of the clustering of awareness of climate change across Nigeria’s 36 states and the Federal Capital Territory (FCT). Third, considering this from a humanitarian health lens provides some insight into the Lancet Countdown on Health and Climate Change in Africa, which called for Perception indicators at the population-level to guide health system adaptation.

The aim of this study is to assess the prevalence of climate change awareness, spatial distribution, sociodemographic factors associated with climate change awareness among adults in Nigeria, explore governance perception of climate change awareness, and discuss the implications for humanitarian health response. The specific objectives are: (1) to determine the national prevalence and sociodemographic distribution of climate change awareness; (2) to examine climate change perceptions such as perceived impact, citizens’ role, government actions urgency, and hazard severity; (3) to assess government performance ratings and responsibility attributions to government for climate change mitigation; (4) to map the spatial distribution of climate indicators across Nigerian 36 states and FCT; (5) to investigate spatial clustering using Global Moran’s I and LISA analysis; and (6) to identify independent predictors of climate change awareness using multivariable logistic regression.

This study contributes to the literature in the following ways. It uses Global Moran’s I and LISA spatial autocorrelation techniques to state-level Afrobarometer data to generate spatially disaggregated results on the clustering of awareness of climate change across Nigeria’s 36 states and the FCT, which were not available before. Secondly, it provides baseline data that is directly relevant to the implementation of Nigeria’s Climate Change Act 2021 and the NDC 3.0 (2025) which calls for measurable and geographically actionable public awareness targets. Thirdly, it highlights a governance accountability deficit in Nigeria, as 76% of the population rated government performance on climate as poor, while 85% said they wanted significantly more action. This is an area that has been under-researched in the peer-reviewed literature on climate awareness in West Africa. Fourth, it explicitly places climate literacy in the context of climate humanitarian health preparedness, thereby linking climate science and humanitarian action in a way that is relevant to Nigeria’s growing burden of floods, droughts and diseases.

## Methods

### Study design and data

Our analysis is based on the Afrobarometer Round 9 Nigeria survey, a nationally representative household survey conducted between 5 and 31 March 2022. Afrobarometer is a pan-African research network that conducts publicly available surveys on governance, democracy, and social development across Africa. The Round 9 Nigeria dataset was chosen because it is the most recent round to include a dedicated climate change module, providing individual-level data on awareness, perceptions, and governance satisfaction that cannot be obtained from any other nationally representative source for Nigeria.

### Study Area and Population

The study was conducted in Nigeria, which comprises 36 states and the Federal Capital Territory (FCT). Nigeria is divided into six geopolitical zones: North-West, North-East, North-Central, South-West, South-East, and South-South. The study population consisted of adults aged 18 years and above who were permanent residents of Nigeria during the survey period.

### Sampling Procedure and Sample Size

Afrobarometer Round 9 utilized a stratified multistage probability sampling design to ensure national representativeness. In the first stage, Enumeration Areas (EAs) were selected as Primary Sampling Units (PSUs) using Probability Proportional to Size (PPS) sampling. Stratification was based on geopolitical zones and place of residence (urban and rural). In the second stage, households within selected PSUs were chosen using a random-walk procedure. In the third stage, one eligible adult respondent was selected from each household using the Kish Grid method to ensure equal probability of selection among household members.

The Afrobarometer Round 9 Nigeria dataset comprises 1,600 respondents, a sample size determined by the Afrobarometer network to be sufficient for nationally representative estimates at the 95% confidence level with a margin of error of approximately 2.5 percentage points. The sample was rounded to 1,600 to improve precision and representativeness across all 36 states and the FCT.

### Study Variables

Primary Outcome Climate change awareness was measured using Afrobarometer item Q67A (“Have you ever heard of the concept of climate change or global warming?”), dichotomized as aware (1) versus unaware (0); respondents selecting “Don’t know” or “Refused” were excluded.

Secondary Outcomes Nine variables captured climate change perceptions and environmental governance, including perceived national impact (Q67B), citizens’ and government roles in mitigation (Q68A–B), drought and flood severity (Q66A–B), government performance on climate change and environmental protection (Q46P–Q), mitigation responsibility (Q69), and adequacy of climate action across stakeholder groups (Q70A–D). Predictor variables include five sociodemographic characteristics which are: sex, age, educational attainment, place of residence and geopolitical zone.

### Statistical Analysis

All analyses were conducted using R statistical software. Categorical variables were summarized using weighted frequencies and percentages. The weighted prevalence of climate change awareness was estimated using:

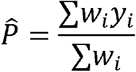

Where:

- 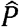 = weighted prevalence
- *W*_*i*_ = survey weight for respondent *i*
- *y*_*i*_= awareness status of respondent *i*

### Bivariate Analysis

Pearson Chi-square (***χ***^2^) tests were used to assess associations between climate change awareness and sociodemographic variables. The chi-square statistic was computed as:

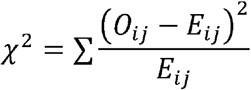

Where:

- *O*_*ij*_= observed frequency
- *E*_*ij*_= expected frequency

Where expected cell counts were less than five, Fisher’s Exact Test was used.

Effect size was measured using Cramér’s V:

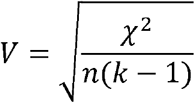

Where:

- *V*= Cramér’s V
- *n*= sample size
- *k*= smaller number of rows or columns

A p-value less than 0.05 was considered statistically significant.

### Logistic Regression Analysis

Binary logistic regression analysis was performed to identify independent predictors of climate change awareness.

The logistic regression model is expressed as:

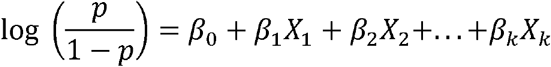

Where:

- *p*= probability of climate change awareness
- *β*_0_= intercept
- *β*_1_…*β*_*k*_= regression coefficients
- *X*_1_… *X*_*k*_ = predictor variables

Adjusted Odds Ratios (AORs) with 95% Confidence Intervals (CIs) were reported.

Model fit was assessed using: Hosmer–Lemeshow goodness-of-fit test, Nagelkerke *R*^2^, Variance Inflation Factors (VIF < 5)

### Spatial Analysis

Spatial analysis was conducted to examine the geographic distribution and clustering of climate change indicators across Nigeria’s 36 states and the Federal Capital Territory (FCT). State administrative boundary shapefiles were obtained from the Global Administrative Areas Database (GADM version 4.1). Individual survey responses were aggregated to produce weighted state-level prevalence estimates.

Choropleth maps were generated using the tmap package in R with quantile classification.

Global spatial autocorrelation was assessed using Moran’s I statistic:

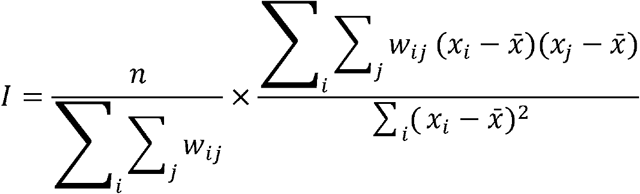

Where:

- *n*= number of spatial units
- *w*_*ij*_= spatial weight between locations *i* and *j*
- *x*_*i*_and *x*_*j*_ = observed values at locations *i* and *j*
- 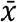 = mean value

Spatial weights were generated using k-nearest neighbors (*k* = 4) with row standardisation.

All spatial analyses were conducted using the sf, spdep, and tmap packages in R.

Ethical Considerations

This study utilized secondary anonymized data obtained from the Afrobarometer Open Data Portal. The original survey obtained informed oral consent from all respondents before data collection. No personal identifiers were included in the dataset, and confidentiality was maintained throughout the analysis. Therefore, additional institutional ethical approval was not required for this study.

## Results

### Socio-demographic Characteristics and Chi-square Analysis

There was a significant awareness gap with fewer than one in three Nigerians saying they had heard of climate change (30.1 per cent, n=438/1,600) (Table 1). Awareness was significantly higher among males than females (35.4% vs. 21.0%; χ^2^=38.6, p<0.001, V=0.158), while older respondents (≥55 years) recorded higher awareness (38.4%) than the youngest age group (26.9%; χ^2^=7.81, p=0.020, V=0.071). Education was the most significant predictor, with awareness increasing significantly from 10.4% in the no education group to 59.1% in the tertiary education group (χ^2^=242.0, p<0.001, V=0.396). Urban residents were more aware than rural counterparts (33.8% vs. 23.9%; χ^2^=17.9, p<0.001, V=0.107), and awareness varied markedly by geopolitical zone, from 17.8% in the North-West to 38.6% in the North-Central zone (χ^2^=51.9, p<0.001, V=0.183).

**Table 1:**
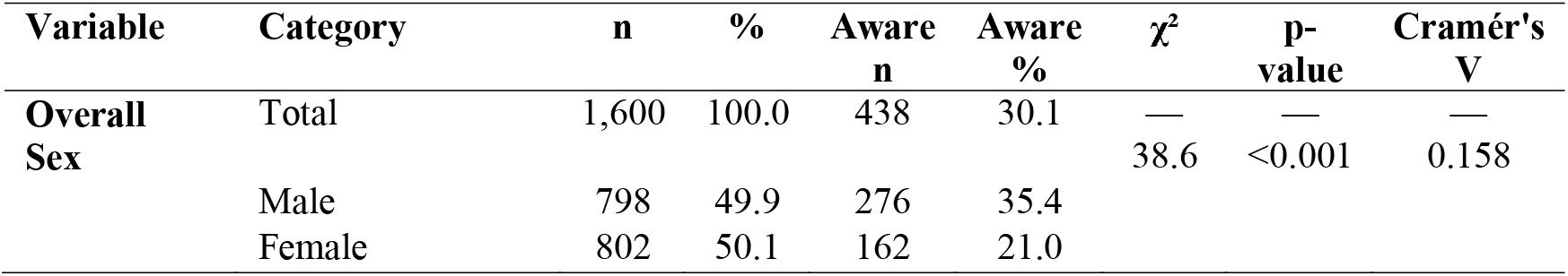
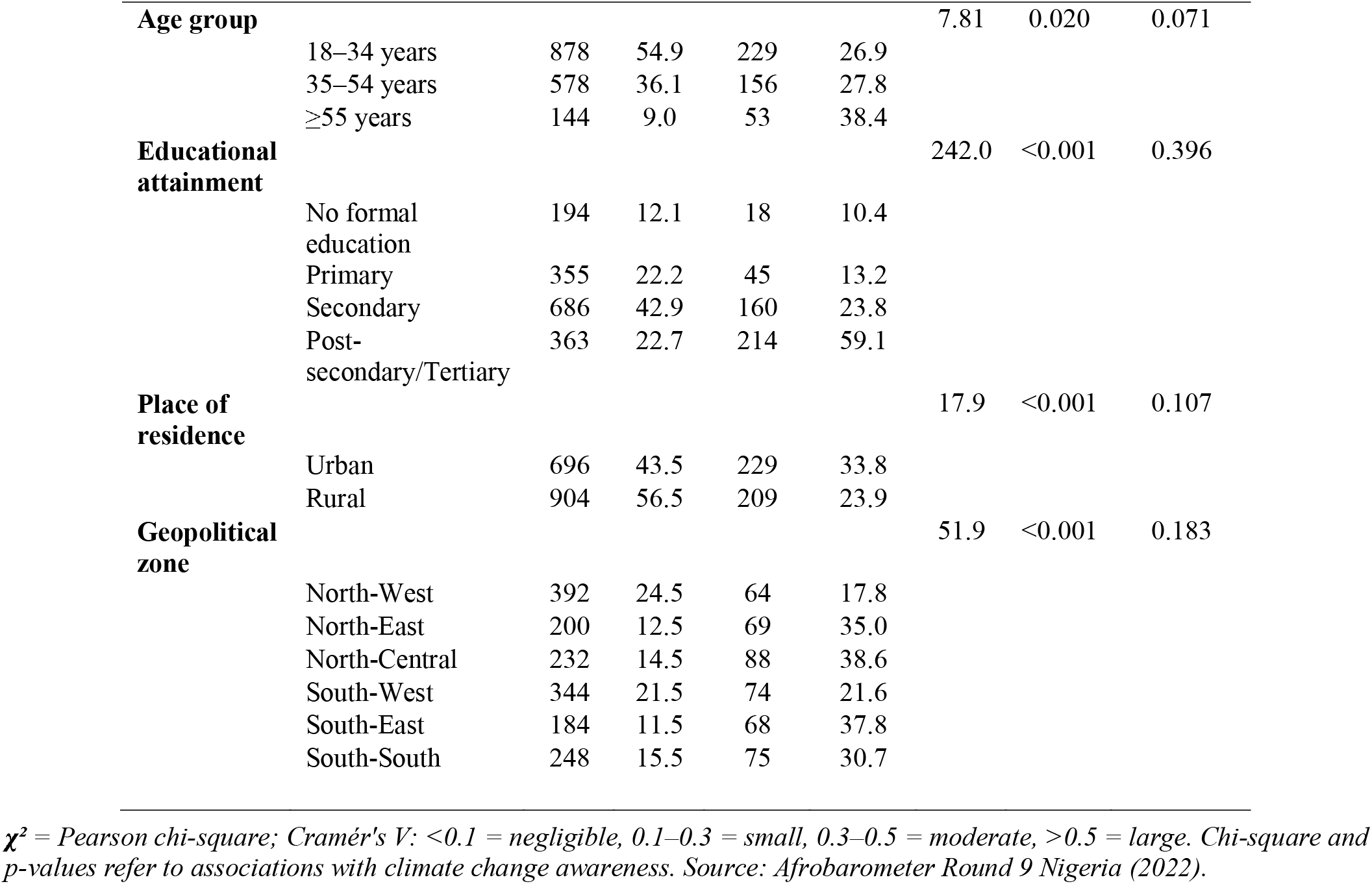
Sociodemographic Characteristics and Prevalence of Climate Change Awareness by Subgroup (n = 1,600)

Education was positively associated with Awareness (Figure 1; χ^2^=242.0, p<0.001, V=0.396) as it rose steadily from 10.4% among those with no formal education to 59.1% among the tertiary graduates, the latter being the only group to exceed the national average of 30.1% (Figure 1). Regionally, North-Central (38.6%), South-East (37.8%), and North-East (35.0%) zones exceeded the national average, while North-West (17.8%) and South-West (21.6%) lagged considerably behind (Figure 2; χ^2^=51.9, p<0.001, V=0.183). The patterns indicate that the least educated and certain northern regions of Nigeria are the least educated, highlighting the need to address climate communication in these groups

**Figure 1.**
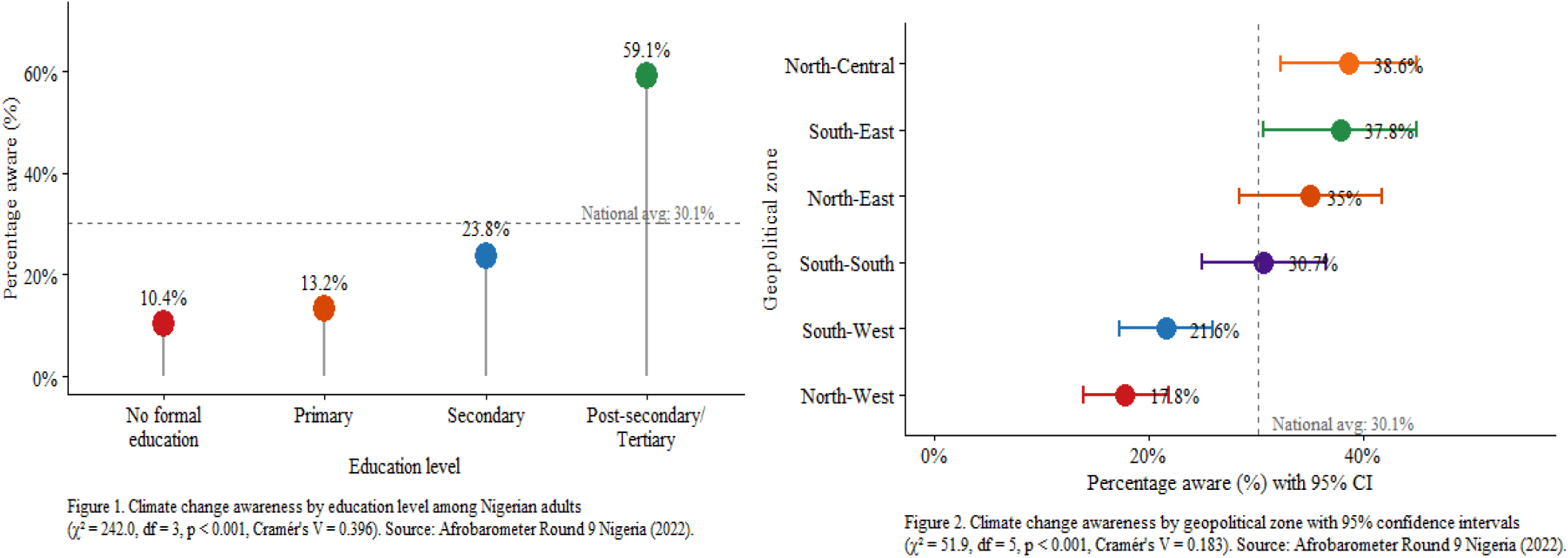
Climate change awareness by education level among Nigerian adults (χ^2^ = 242.0, df = 3, p < 0.001, Cramér’s V = 0.396). Source: Afrobarometer Round 9 Nigeria (2022).

According to table 2, the majority of the climate-change-aware respondents felt that climate change is making Nigeria’s conditions worse (64.6%), that the citizens have a role to play in curbing climate change (74.5%) and that the government should respond urgently (71.9%). The association between drought hazard severity and awareness was negligible, with aware respondents more likely to think that droughts are worsening than unaware respondents (29.3% vs. 26.0%; χ^2^=8.0, p=0.018, V=0.074). Flooding severity did not differ significantly between groups (24.4% vs. 24.7%; χ^2^=5.89, p=0.053, V=0.063). Widespread public dissatisfaction with government’s climate and environmental governance in Nigeria was evident in the low ratings given to government performance on both climate change (3 in 4 rated it badly; 76.0%) and environmental protection (nearly as many; 71.9%, Table 2).

**Table 2:**
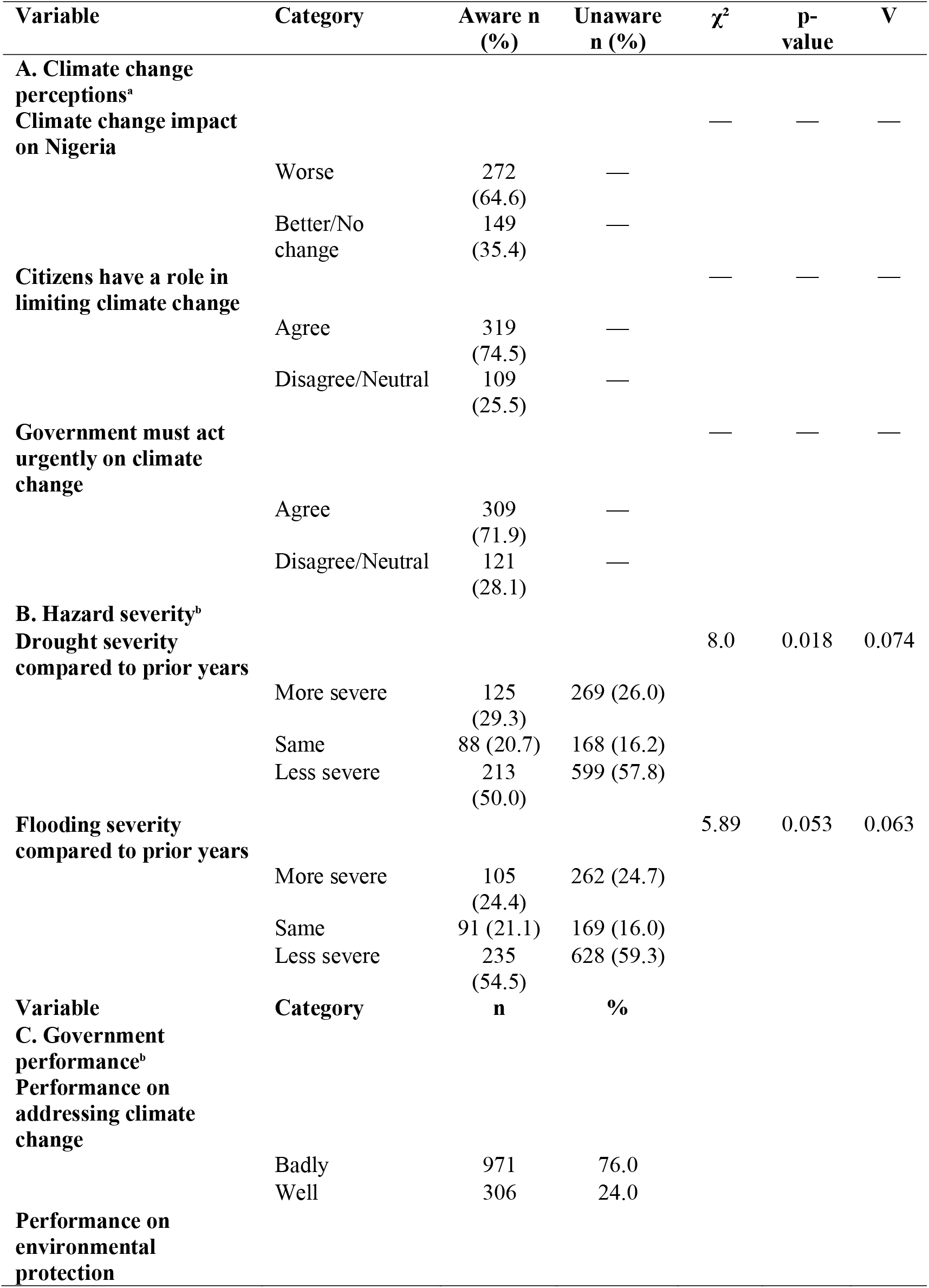
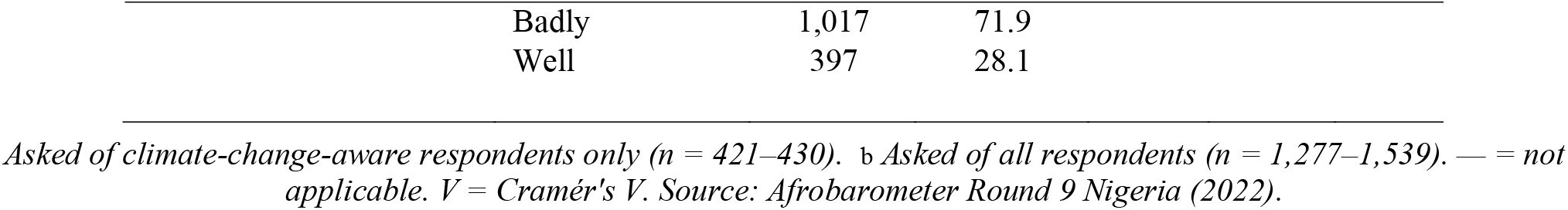
Climate Change Perceptions, Hazard Severity, and Government Performance Ratings.

| Variable | Category | Aware n (%) | Unaware n (%) | $\chi^2$ | p-value | V |
| --- | --- | --- | --- | --- | --- | --- |
| **A. Climate change perceptions<sup>a</sup>** |  |  |  |  |  |  |
| Climate change impact on Nigeria |  |  |  | — | — | — |
|  | Worse | 272 (64.6) | — |  |  |  |
|  | Better/No change | 149 (35.4) | — |  |  |  |
| **Citizens have a role in limiting climate change** |  |  |  | — | — | — |
|  | Agree | 319 (74.5) | — |  |  |  |
|  | Disagree/Neutral | 109 (25.5) | — |  |  |  |
| **Government must act urgently on climate change** |  |  |  | — | — | — |
|  | Agree | 309 (71.9) | — |  |  |  |
|  | Disagree/Neutral | 121 (28.1) | — |  |  |  |
| **B. Hazard severity<sup>b</sup>** |  |  |  |  |  |  |
| **Drought severity compared to prior years** |  |  |  | 8.0 | 0.018 | 0.074 |
|  | More severe | 125 (29.3) | 269 (26.0) |  |  |  |
|  | Same | 88 (20.7) | 168 (16.2) |  |  |  |
|  | Less severe | 213 (50.0) | 599 (57.8) |  |  |  |
| **Flooding severity compared to prior years** |  |  |  | 5.89 | 0.053 | 0.063 |
|  | More severe | 105 (24.4) | 262 (24.7) |  |  |  |
|  | Same | 91 (21.1) | 169 (16.0) |  |  |  |
|  | Less severe | 235 (54.5) | 628 (59.3) |  |  |  |
| **Variable** | **Category** | **n** | **%** |  |  |  |
| **C. Government performance<sup>b</sup>** |  |  |  |  |  |  |
| **Performance on addressing climate change** |  |  |  |  |  |  |
|  | Badly | 971 | 76.0 |  |  |  |
|  | Well | 306 | 24.0 |  |  |  |
| **Performance on environmental protection** |  |  |  |  |  |  |

|  |  |  |
| --- | --- | --- |
| Badly | 1,017 | 71.9 |
| Well | 397 | 28.1 |

*Asked of climate-change-aware respondents only (n = 421–430). <sup>b</sup> Asked of all respondents (n = 1,277–1,539). — = not applicable. V = Cramér's V. Source: Afrobarometer Round 9 Nigeria (2022).*

Multivariable adjusted independent factors of climate change awareness are shown in Table 3. Males had 67% higher odds of awareness than females (AOR=1.67, 95% CI: 1.29–2.16, p<0.001), and respondents aged ≥55 years were nearly twice as likely to be aware as the youngest group (AOR=1.76, 95% CI: 1.13–2.74, p=0.012). The strongest predictor was education – those who received a tertiary education were over ten times more likely to be aware of the warnings than were those with no education (AOR=10.44, 95% CI: 5.97–19.12, p<0.001), while the odds of being aware were 36% higher for those living in urban areas than for those living in rural areas (AOR=1.36, 95% CI: 1.04–1.79, p=0.026). The North-East residents were significantly more aware than North-West counterparts with an AOR of 2.06, and the North-Central residents were also significantly more aware with an AOR of 1.72 as opposed to the South-West counterparts who had an AOR of 0.58; p=0.014. The model had a goodness of fit (Hosmer-Lemeshow χ^2^=5.96, df=8, p=0.652) and explained 25.5% of the variance in climate change awareness (Nagelkerke R^2^=0.255); no multicollinearity between the predictors was observed, with all VIF values below 5.

**Table 3:**
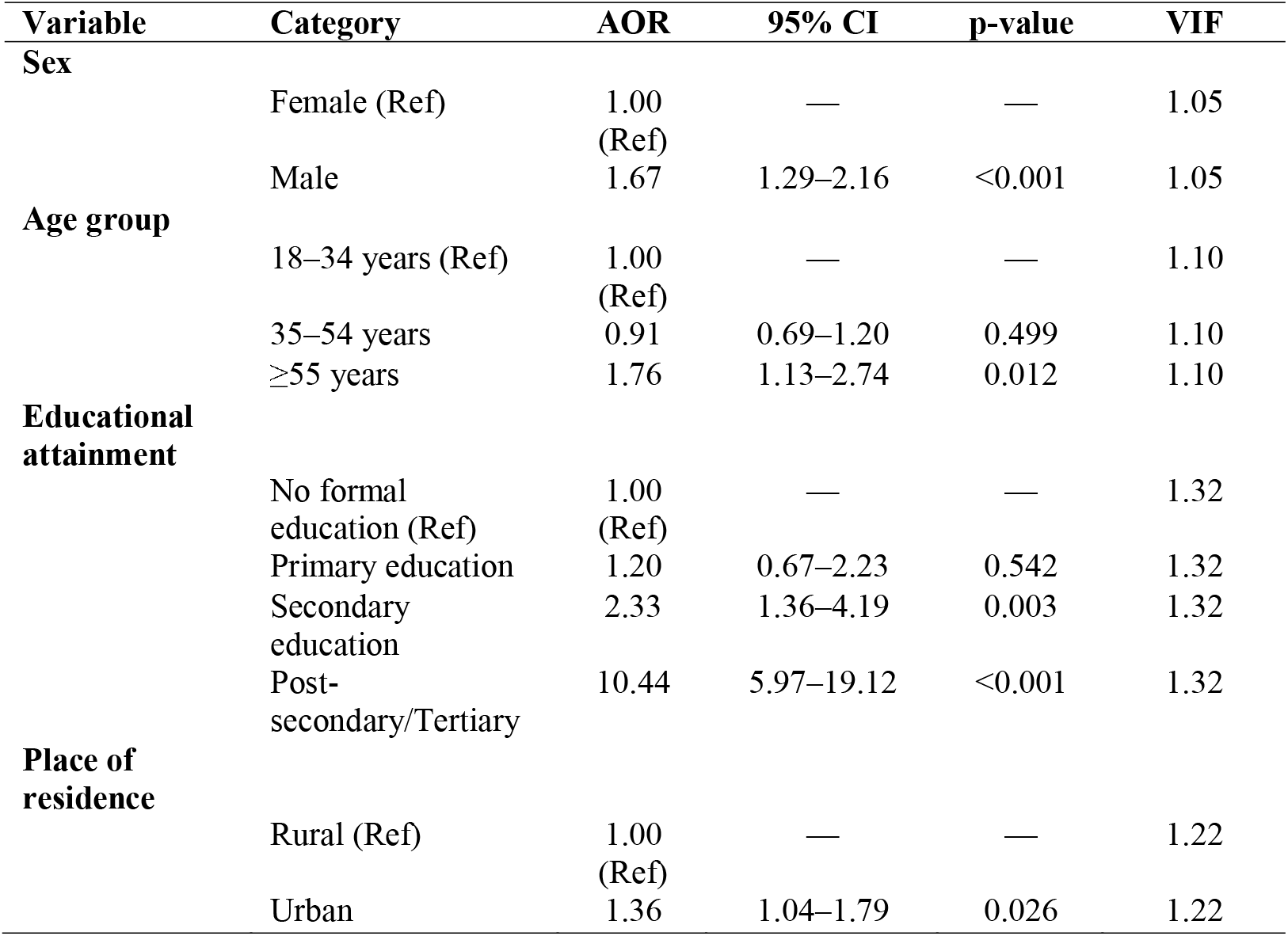
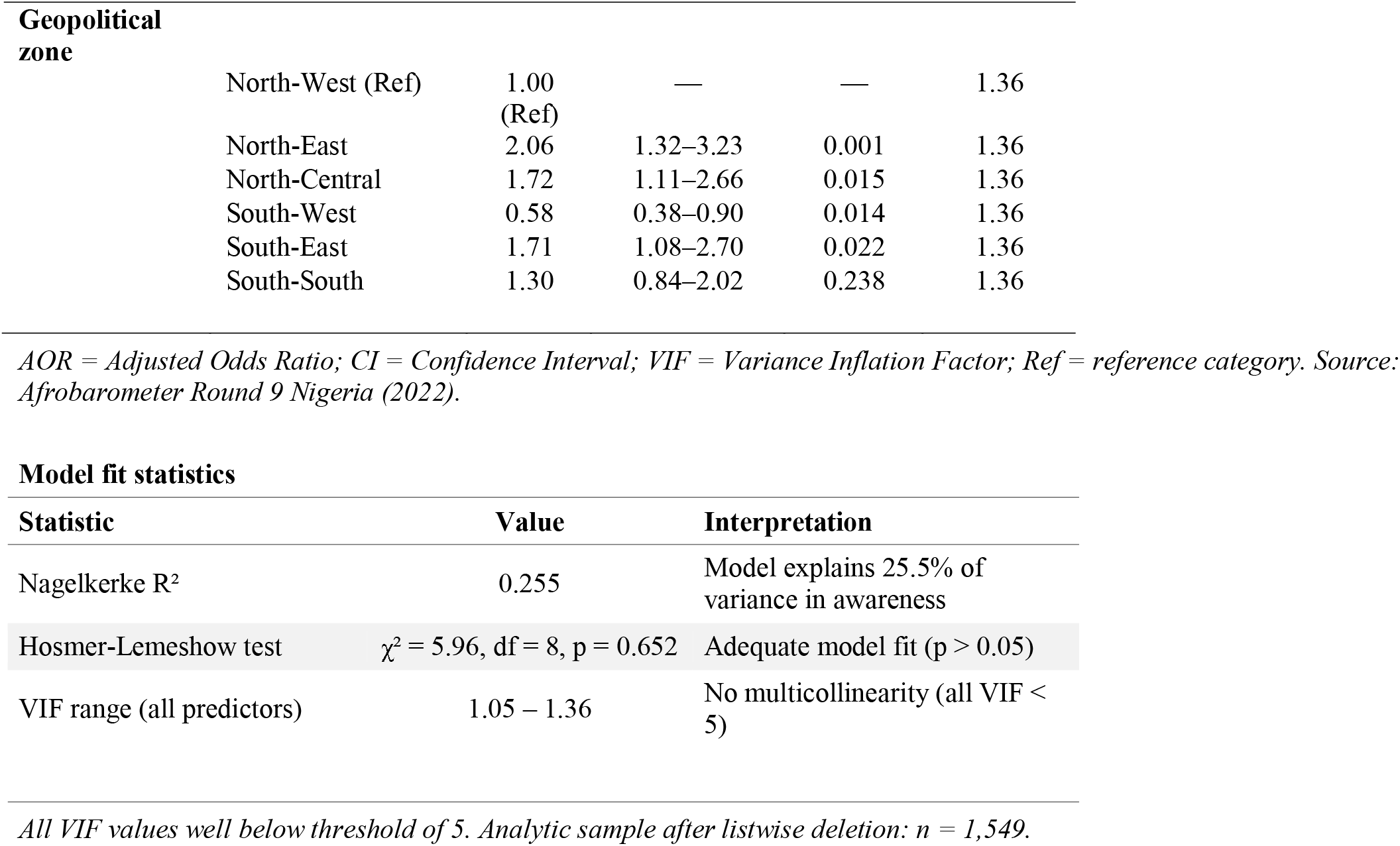
Multivariable Logistic Regression: Independent Predictors of Climate Change Awareness (n = 1,549)

### Model fit statistics

The spatial distribution of climate change indicators was shown in Table 4 by states of Nigeria. Global Moran’s I statistics showed that there is significant positive spatial autocorrelation for climate change awareness (I=0.199, z=2.275, p=0.011), climate change drought severity perception (I=0.162, z=1.876, p=0.030) and climate change government performance (I=0.470, z=4.834, p<0.001), suggesting that states in close geographic proximity have similar values for these indicators. There was no significant spatial clustering of severity of flooding (I=−0.073, p=0.676) indicating a random distribution of flooding by state. Localized spatial patterns of substantive public health interest were detected using LISA cluster analysis. For climate change awareness, the states of Nasarawa (30.4%) were situated in the high-high cluster while Taraba was a low-low cold spot (25.0%) with low awareness states flanked by relatively high awareness states; this low-high combination of low awareness surrounded by relatively high awareness states was referred to as low-high outliers, and these states were identified as Kebbi (31.3%), Sokoto (31.9%) and Zamfara (32.7%). The drought severity hot spots were mostly in the North-East which had a higher proportion of 46.9%, 31.2% and 41.7% for Adamawa, Borno and Gombe respectively, which is in line with the region’s documented vulnerability to drought. The highest clustering was for the government climate ratings across the south-east states with all states in this region forming a clear high-high cluster (84.2%–96.4%), which suggests that the government performance ratings in the region are consistently poor, whilst the northern states of Kaduna, Katsina and Sokoto were identified as low-high outliers (57.5%–71.9%), indicating that the government performance ratings in the region were relatively better, but still low.

**Table 4:**
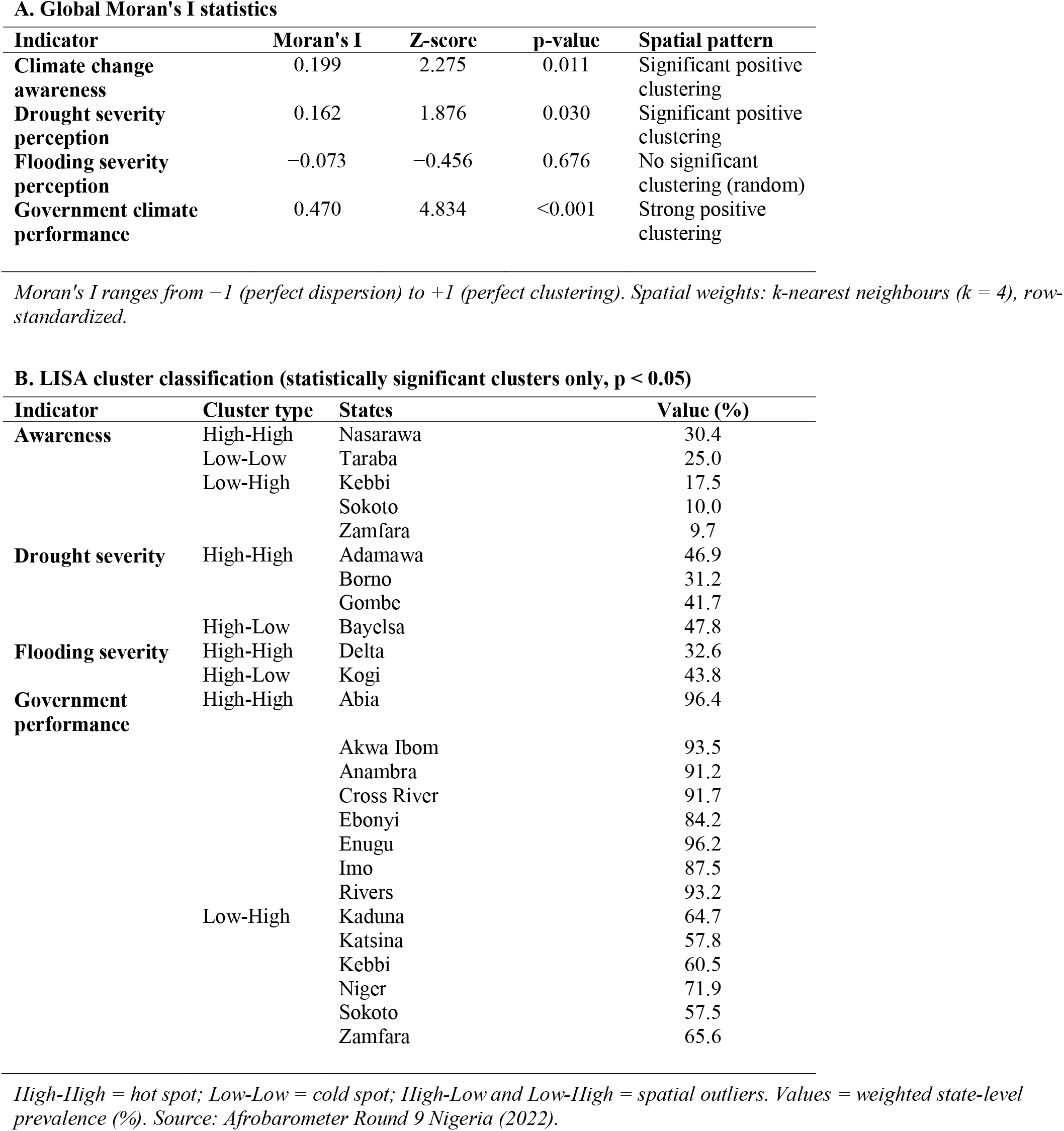
Global Moran’s I Spatial Autocorrelation Statistics and LISA Cluster Classification of Climate Change Indicators Across Nigerian States.

The choropleth maps provide a compelling geographic characterization of climate change knowledge, hazard awareness and climate governance in Nigeria. The North-Central and South-Eastern states are visibly distinct with the bulk of the states showing deep blue color indicating high levels of awareness (43.48–87.50%), while the North-West states show the lightest colors (0.00–15.35%) reflecting the low levels of awareness, and this spatial delineation is a clear representation of the inequalities in educational and infrastructural provision across Nigeria. The perceptions of drought severity (Figure 3B) do not match this pattern, with the darkest browns in the North-East (39.82– 62.50%), an area that is a desertification hotspot and saddled with the encroaching Sahara — a perception that is not only subjective but rooted in the ecological realities of the region. The severity of flooding (Figure 3C) shows a more complex spatial pattern, with teal color patches found in the South-Western and North-Central areas (34.40 - 66.70%). This reflects the hydrodynamic complexity of river basins and coastal lowlands in Nigeria. Most distinctly, figure 3D shows a public verdict on government climate performance that is almost universally negative, with the deep crimson covering the South-Eastern states (91.60-96.40%) being an almost complete blanket of public dissatisfaction with the quality of governance, and even the lightest shade in the North-West (46.20-61.11%) showing majority discontent with the quality of governmental climate action.

**Figure 3.**
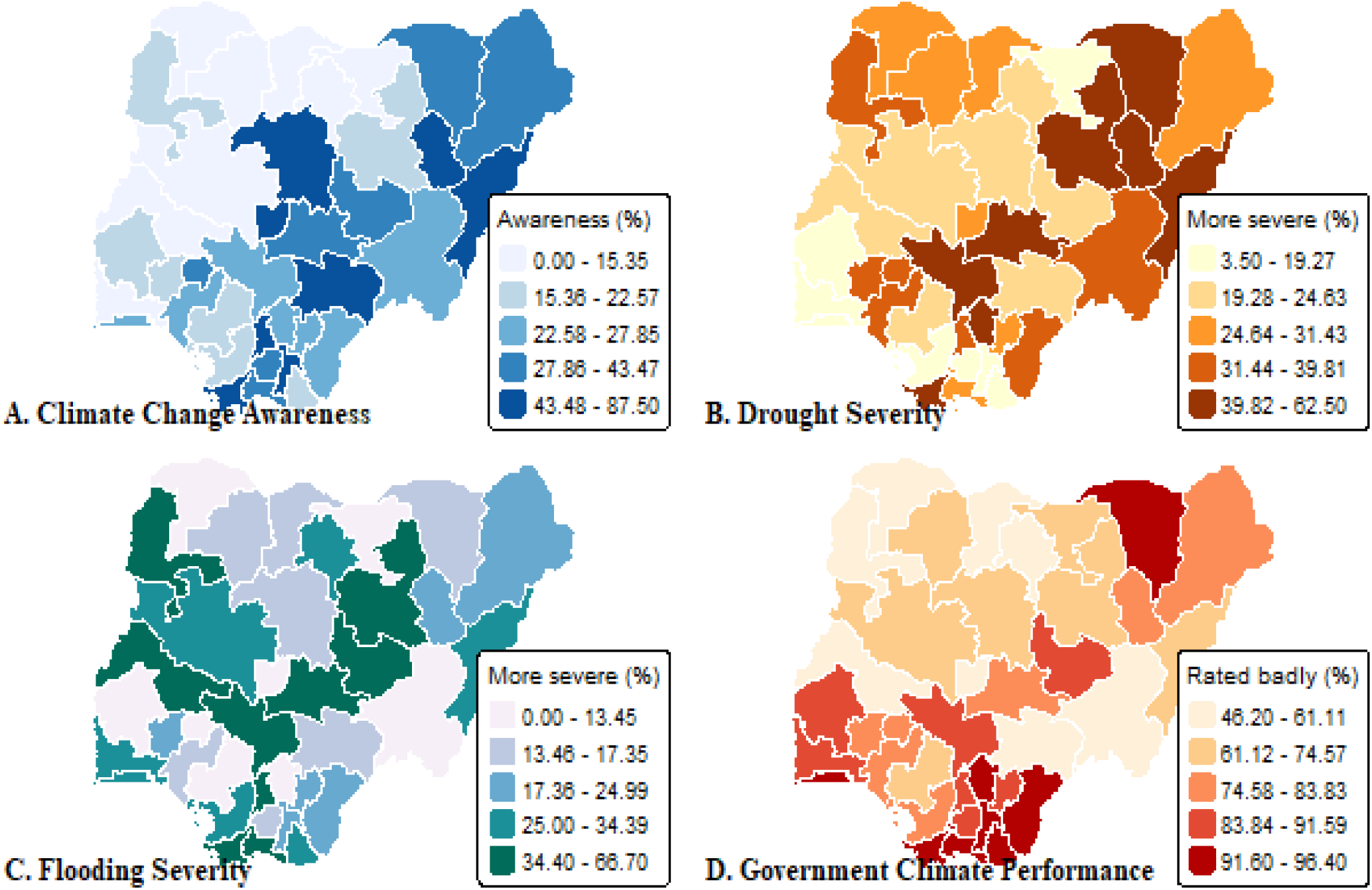
Spatial distribution of climate change awareness (A), drought severity perception (B), flooding severity perception (C), and government climate performance ratings (D) across Nigerian states. Source: Afrobarometer Round 9 Nigeria (2022)

When climate change-aware respondents were asked who was mainly responsible for controlling climate change, the Nigerian government was overwhelmingly cited (76.5%), followed by ordinary citizens (13.5%) and developed countries (3.7%) (Table 5). In every stakeholder group, a clear lack of satisfaction was expressed regarding the current efforts of the government, business and industry, developed countries and ordinary citizens, with 85.0% of people expressing dissatisfaction with government actions, followed by business and industry with 73.5%, and developed countries with 76.7% and ordinary citizens with 57.5%. Notably, less than 1 in 10 respondents felt that any group was doing enough, underlining the deep and pervasive feeling that action on climate is sorely lacking at all levels.

**Tabl5 5:** Primary Responsibility Attribution and Stakeholder Effort Sufficiency Among Climate-Change-Aware Respondents.

| Variable | Category | n | % |
| --- | --- | --- | --- |
| Primary responsibility for |  |  |  |
| **limiting climate change (n = 429)** |  |  |  |
|  | Government of Nigeria | 328 | 76.5 |
|  | Ordinary citizens | 58 | 13.5 |
|  | Developed countries | 16 | 3.7 |
|  | No one | 12 | 2.8 |
|  | Business and industry | 11 | 2.6 |
|  | Traditional leaders | 4 | 0.9 |
| **Are stakeholders doing enough to limit climate change?** |  |  |  |
| **Ordinary citizens (n = 416)** |  |  |  |
|  | Need a lot more | 239 | 57.5 |
|  | Need somewhat more | 141 | 33.9 |
|  | Doing enough | 36 | 8.7 |
| **Business and industry (n = 422)** |  |  |  |
|  | Need a lot more | 310 | 73.5 |
|  | Need somewhat more | 104 | 24.6 |
|  | Doing enough | 8 | 1.9 |
| **Government of Nigeria (n = 426)** |  |  |  |
|  | Need a lot more | 362 | 85.0 |
|  | Need somewhat more | 55 | 12.9 |
|  | Doing enough | 9 | 2.1 |
| **Developed countries (n = 407)** |  |  |  |
|  | Need a lot more | 312 | 76.7 |
|  | Need somewhat more | 74 | 18.2 |
|  | Doing enough | 21 | 5.2 |

*All variables asked of climate-change-aware respondents only. Source: Afrobarometer Round 9 Nigeria (2022).*

## Discussion

In this study, less than one-third (30.1%) of Nigerian adults had heard of climate change, a relatively low figure compared to 56% across the continent and the lowest level found in 38 African countries surveyed (Afrobarometer, 2025). This figure is a reflection of a structural lack of knowledge, caused by educational exclusion, gender inequality and geographic marginalization, which intersect with Nigeria’s humanitarian fault lines, making the country the 154th most vulnerable to climate change of 185 countries in the ND-GAIN climate vulnerability index, and ranking 2nd in the UNICEF Children’s Climate Risk Index (2021, 2022; van der Linden, 2015).

The strongest independent predictor, by far, was education (AOR=10.44; 95% CI: 5.97–19.12), which aligns with cross-national evidence that education is the most powerful predictor of climate literacy (Lee et al., 2015; Simpson et al., 2021). Awareness was lower than the national average among only tertiary graduates, falling to 10.4% among those with no formal schooling. This means that climate knowledge in Nigeria is structurally created through formal education; individuals who do not come from the formal education track are systematically excluded from climate discourse, and climate awareness campaigns geared toward the general public will have limited impact on populations which lack the basic capacity and resources for receiving and processing climate information. This is supported by the gender gap. Gender differences in climate awareness persisted, with males being 67% more likely to be climate-aware than females (AOR=1.67; p<0.001), highlighting disproportionate access to education and media, as well as early warnings, which is a clear disadvantage to rural women, the same group facing the highest burden of food insecurity, displacement, and infectious diseases due to climate change (Adeola et al., 2024).

The odds of awareness were 36% greater for urban residents than rural residents (AOR=1.36; p=0.026); higher likelihood of awareness among older respondents (≥55 years; AOR=1.76; p=0.012) may be attributable to increased lived experience of environmental change but not necessarily formal information access (González & Sánchez, 2022). However, residents in the North-East and North-Central region were significantly more aware than those in the North-West region (AOR=2.06; p=0.001 and AOR=1.72; p=0.015 respectively) suggesting a difference in educational infrastructure and media penetration across the geopolitical zones (Azeez et al., 2024). The results of spatial analysis were positive which showed that there was no randomness in geopolitical variation of awareness.

The result of Global Moran’s I showed significant positive clustering for awareness (I=0.199, p=0.011), indicating that low-awareness states clustering together with other low-awareness states. Based on LISA analysis, the most critical states for geographically differentiated intervention are low-high spatial outliers, with Kebbi (17.5%), Sokoto (10.0%) and Zamfara (9.7%) having low awareness levels despite being surrounded by states with higher awareness levels. The perception of drought severity was separated into hotspot areas in Adamawa (46.9%), Borno (31.2%) and Gombe (41.7%) — states that experienced active conflict, displacement and the ecological effects of shrinking Lake Chad (United Nations Security Council, 2024). In these populations, droughts are being manifested with ecological fidelity but not accompanied by the mental model of climate change needed to give it a name, to make decisions in response to it, or to hold parties accountable for climate action. The same is true with regard to government climate performance, which exhibited the highest spatial clustering of any indicator (I=0.470, p<0.001), with all 8 states in the South-Eastern cluster of poor climate performance ratings, which included Enugu (96.2%), Abia (96.4%), Cross River (91.7%) and Rivers (93.2%). This concentration cannot be attributed to lack of awareness as above average climate literacy is reported in the South-Eastern states.

Low awareness and governance failure are therefore border phenomena, where one in one area is not the same as the other in another area, and such an amalgamation under a single national response would be a false economy and lead to both challenges not being adequately met. If populations feel responsible for government for the primary responsibility for climate change but find it to be too unresponsive, it undermines the institutional trust that underpins effective humanitarian health coordination (Torsu & Kronke, 2025; Duntoye et al., 2023). The humanitarian health impacts of these gaps are well known. The 2022 floods in Nigeria killed 603 persons and displaced 1.4 million people while the 2024 floods displaced more than 5.2 million Nigerians (NEMA, 2023; NEMA, 2025). Kebbi, Sokoto, and Zamfara are the three low-awareness spatial outliers that are located in an area of growing malaria transmission, frequent displacement by floods, and rapid desertification. Lacking climate literacy, these communities will not be able to attribute worsening disease conditions to climate change, will be unable to adopt protective health behaviors in response, and will not be able to meaningfully engage in early warning systems that humanitarian health response relies upon (van der Linden, 2015; WHO, 2023).

Governance discontent in the South-East is over 90%, and that lowers the effectiveness of top-down preparedness interventions, no matter how technically sounds the interventions themselves are. The North-West is vulnerable to humanitarian crises due to low awareness, and the South-East is vulnerable because of governance failures; these two pathways to humanitarian health vulnerability require two different kinds of responses. These results confirm that climate awareness is a structural deficit, with a concentration of the deficit at the spatial level and different mechanisms of operation across Nigeria. To bridge this gap, significant attention must be paid to the need for both spatially differentiated and gender-responsive, and education-based programming as part of the implementation mechanism of Nigeria’s Climate Change Act 2021 and NDC 3.0 commitments (Federal Republic of Nigeria, 2021; Federal Republic of Nigeria, 2025).

Humanitarian health actors need to make climate literacy part of their flood/drought response programming, as communities who know nothing about the structural cause of the hazards they face cannot make informed choices for adaptation. To break the link between educational exclusion and climate illiteracy across generations, gender-transformative strategies need to be implemented using community radio, local-language broadcasting, and faith-based approaches, and these strategies must be formally incorporated into national school curricula (Lee et al., 2015; Simpson et al., 2021). More studies are needed to apply longitudinal designs and explore if the increasing intensity of climate shocks in Nigeria, such as the floods in 2024 that impacted 5.2 million people, have altered public perceptions and governance assessments, which this cross-sectional design cannot capture (NEMA, 2025). The potential to track changes over time would be provided by a comparative analysis leveraging data from the Afrobarometer Round 10 Nigeria survey, revealing whether increased exposure to climate shocks (2022-2024) has significantly changed perceptions and governance assessments.

## Data Availability

Data Availability: All data used in this study are publicly available. The Afrobarometer Round 9 Nigeria dataset can be freely accessed and downloaded from the Afrobarometer website at www.afrobarometer.org. No additional or restricted datasets were used in this study, and no new data were generated. The data are available to any researcher wishing to replicate or build upon the findings of this study.

https://www.afrobarometer.org/survey-resource/nigeria-round-9-data-2023/

